# Analysis of Burkina Faso’s institutional framework and budget allocations for NCDs control and monitoring

**DOI:** 10.1101/2024.10.29.24316362

**Authors:** Moussa Ouedraogo, Dia Sanou, Mahamadé Goubgou, Estelle Aissa Bambara, Souleymane Tirogo, Aly Savadogo

**Affiliations:** Doctoral School of Sciences and Technology, Laboratory of Applied Biochemistry and Immunology (LABIA), Joseph Ki-ZERBO University, Ouagadougou, Burkina Faso; Nutrition Directorate, Ministry of Health, Ouagadougou, Burkina Faso; FAO, Deputy Country Representative, Bangladesh; National Center for Scientific and Technological Research (CNRST)/Health Sciences Research Institute (IRSS), Burkina Faso

**Keywords:** governance, institutional framework, financial allocation, non-communicable diseases

## Abstract

Diet-related non-communicable diseases (NCDs) are today a major public health problem and a global development challenge. Yet governance responses to these diseases are still in their infancy in most low-income countries like Burkina Faso. How Burkina Faso organizes itself institutionally and financially to respond adequately to NCDs is unknown to the scientific community. The aim was to analyze Burkina Faso’s institutional framework and budget allocations. This was a cross-sectional survey based on mixed (qualitative and quantitative) data collection. The analysis revealed a number of difficulties in the institutional framework hindering the performance of the fight against NCDs in Burkina Faso, including (i) the absence of a multi-sectoral policy or strategic plan involving all stakeholders, (ii) the absence of a multi-sectoral coordinating body and (iii) the lack of sufficient financial resources. A total of twentynine (29) budget lines related to the prevention and/or management of NCDs were identified, with a total budget of 17.33 billion FCFA ($29.8 million), or an average of $2.72 million per year. This represents only 1.55% of the total budget of the Ministry of Health over the same period. We recommend, among other things, the development of a national multi-sectoral policy with a clear definition of the roles and responsibilities of each player, the creation of a coordinating body, improved funding, and greater attention to NCDs in the provision of primary healthcare services.

## 1. Introduction

Diet-related non-communicable diseases (NCDs) are today a major public health issue and a global development challenge [1]. Also known as chronic diseases, they include obesity, cardiovascular disease, diabetes, cancer, hypertension, hyperglycemia and oral diseases [2]. According to the Global Panel’s experts, the health risks of chronic food-borne diseases are greater than the combined risks of tobacco, alcohol and unprotected sex [3]. NCDs account for over 50% of total premature mortality (i.e. deaths under the age of 60) in most low- and middle-income countries [4, 5]. This is compounded by insufficient budgetary allocations to the fight against NCDs over the last ten years [6]. According to the results of STEPS surveys in Burkina Faso, the prevalence of obesity (BMI>30kg/m^2^) rose from 11.3% in 2013 to 15.9% in 2021 in urban areas [7, 8] and WHO estimates that over 35% of deaths in Burkina Faso are due to NCDs [9]. The cost of inaction far outweighs the cost of action against non-communicable diseases, as recommended in the global action plan proposed by the WHO [10]. However, it is encouraging to note that several global initiatives have been launched to tackle the global burden of NCDs [11]. The World Health Assembly has adopted a global action plan 2013-2020 on NCDs that calls for country-level capacity building, leadership, governance, multi-sectoral action and partnerships to accelerate the fight against NCDs in countries [12]. Despite international interest in the fight against NCDs, the response in most low- and middle-income countries is still in its infancy [13, 14]. Yet with the rapid socio-economic transitions taking place in sub-Saharan Africa, if nothing is done, there is a risk that the growing prevalence of NCDs will overwhelm already struggling health services, with adverse consequences for individuals and economies [2, 15]. WHO estimates that NCDs will be the leading cause of death in sub-Saharan Africa by 2030 [16]. Ouedraogo et al. showed that many concerns about the governance of NCDs remain unresolved in most ECOWAS countries, namely the question of institutional arrangements and funding for NCD control [14]. The aim was to analyze Burkina Faso’s institutional framework and budget allocations for NCD control.

## 2. Methodology

The theoretical approach is based on Ground Theory (GT) [17]. GT proposes an approach that favors analysis in data from several sources, in order to triangulate the information collected. The central principle in data analysis is the constant return to comparison between the products of analysis and empirical data [17]. On the one hand, it was a cross-sectional study based on the mixed (quantitative and qualitative) data collection proposed by Halcomb et al,[18] with key informants, combined with a literature review and an analysis of budget allocations for the fight against NCDs. This second part is based on the methodological approach developed by the Scaling Up Nutrition Movement [19], adapted by UNICEF [20]. This methodology, developed for the purpose of monitoring investments in nutrition, has been adapted to NCDs within the framework of this study. Only the budget allocations of the Ministry of Health were analyzed, due to the difficulty of obtaining accurate data on funding from technical and financial partners. The budget lines taken into account in this second phase are those linked to the prevention and control of NCDs.

### 2.1. Target population

The survey mainly involved resource persons from the Ministry of Health, identified on the basis of a review of the 2016-2020 integrated strategic plan to combat NCDs (PILMNT) [21]. These people have been identified on the basis of the involvement of the structure for which they are responsible in implementing the plan.

The document review consisted in examining reference documents (policies, strategic plans and programs) relating to NCDs in order to triangulate information. A total of 20 interviews were conducted with (15) technical structures of the Ministry of Health and Public Hygiene and five (5) NGOs/Associations working in the health sector. Interviewees had to (i) be in charge of their structure (or have been designated by the person in charge), (ii) have given informed consent to take part in the study, and (iii) have been involved in the project.

### 2.2. Tool and data collection

A semi-structured questionnaire built around the components of NTM governance proposed by Ouedraogo et al. notably (i) involvement in the elaboration of reference documents, notably the PILMNT, (ii) multi-sectoral collaboration, (iii) availability of a common results framework, (iv) existence of a consultation framework, (v) availability of sufficient resources for the implementation of national interventions. Interviews were recorded with a Dictaphone and partially transcribed for analysis purposes. Data collection took place from January to March 2023.

With regard to budget tracking, the analysis focused on the final allocations and expenditures of the Burkina Faso Ministry of Health over the period 2010 to 2020. These data were retrieved from the Integrated Expenditure Circuit (CID) platform of the Ministry of Economy, Finance and Development (MINEFID). The platform groups together all operations containing allocations and expenditures, as well as transfers made by the government to local authorities and other public establishments. At the request of the research team, data from 2010 to 2020 were extracted from the CID by a MINEFID agent, containing allocations and expenditures, and then made available to the authors for the various analyses. The authors then carried out a detailed examination of the Ministry of Health’s budget to extract the budget lines relating to NCDs.

Certain budget lines have not been taken into account in line with the SUN approach used. Indeed, the methodology suggests that budget lines should not be included in the analysis when they concern:

- the payment of civil servants’ salaries;
- the operation of general and technical services; hospitals, health districts, training establishments, etc;
- the organization of examinations and competitions;
- operating and project expenditure.

The research team then analyzed the budget lines selected during the data collection phase in order to categorize and weight them. Categorization made it possible to classify the selected budget lines into 3 categories: specific, sensitive and positive (Figure 1). The validated budget lines were then weighted. This involved assigning a rate to each line according to its estimated level of contribution to the prevention and control of NCDs in the country. This rate was determined by the authors on the basis of the available scientific evidence, the current state of the intervention implemented in the country, the context of NCDs at national and international level, and finally on the basis of the interventions proposed in the Action Plan for the Prevention and Control of Non-Communicable Diseases 2013-2020 [10, 22]. Budget lines considered specific to NTMs were given a weighting of 100%. As for those classified as sensitive to NTMs, three levels of weighting were applied according to the estimated degree of sensitivity of the investments, i.e. 10%, 25% and 50% for low, medium and high sensitivity investments respectively. Favorable investments were not included in total NTM expenditure. Consequently, a zero rate has been applied to these lines.

**Figure 1.**
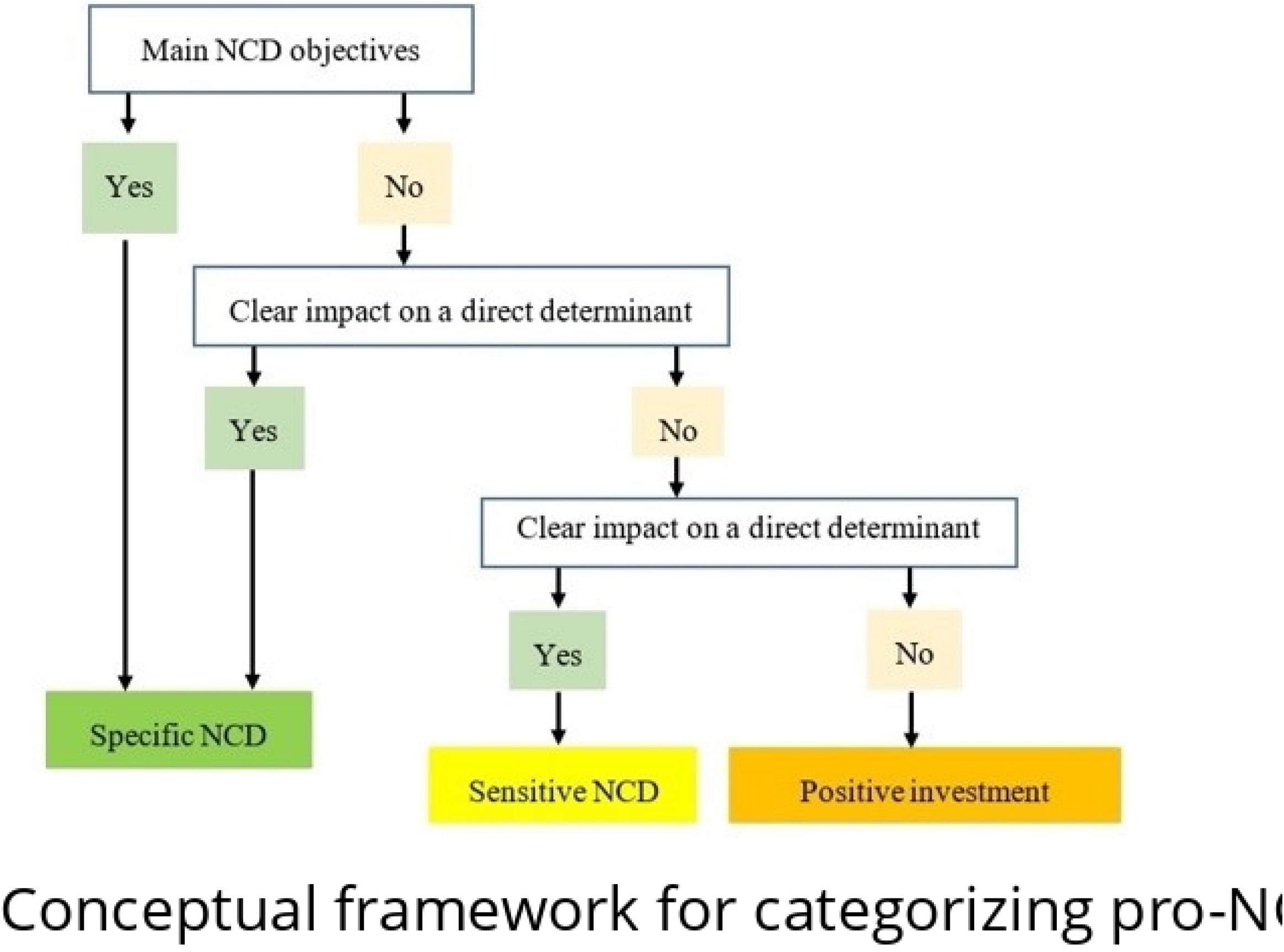
Conceptual framework for categorizing pro-NCD budget lines adapted from the conceptual framework for categorizing pro-Nutrition budget lines.

### 2.3. Ethical consideration

The study was approved by the Ethics Committee for Health Research (CERS) based in Ouagadougou, Burkina Faso in its deliberation no. 2021-03-049 2021. All participants interviewed as part of this study were interviewed after giving their verbal consent.

### 2.4. Data processing and analysis

An analysis matrix was designed, then filled in from the various transcripts. The matrix was then analyzed by thematic group. This analysis concerns only actors from the Ministry of Health. The level of collaboration was assessed primarily on the basis of participants’ statements. Verbatims were used to support the analyses.

For the analysis of budget allocations, the database obtained at the end of the various stages was processed using Stata 12 software. The data were then subjected to descriptive analysis and calculations of budget allocation frequencies. Trends in budget allocations were examined over time. Observed differences were evaluated using Student’s t-test, with a significance level of 5% and a confidence interval of 95%.

## 3. Results

### 3.1. Institutional arrangements

The literature review, and in particular an examination of the organizational chart of the Ministry of Health and Public Hygiene, shows that Burkina Faso has set up a Department for the Prevention and Control of Non-Communicable Diseases (DPCM) within the General Directorate of Health and Public Hygiene, to better coordinate all NCD control programs and pool energies at national, sub-regional and international levels (Figure 2). Alongside this technical department, there are other departments whose missions directly or indirectly include the fight against NCDs. This is the case of the Nutrition Directorate, for example, which implements the multisectoral National Nutrition Policy 2020 - 2029, one of whose major strategic axes is “Strengthening the fight against overnutrition and nutrition-related chronic non-transmissible diseases” (Strategic Axis III) [23]. Although these structures report to the same general management, they seem to function mainly as technical units, rather than as specific program management and implementation entities. Indeed, it emerged from the various interviews that each structure had its own action plan and annual work plan, and that the preparation of these documents was in most cases carried out in a compartmentalized manner. “*…*.. *If not, as long as each department works in isolation from the other, it will be difficult to achieve the expected results, as each department will pursue its own agenda rather than the common agenda of the general management…*” Alongside these technical directorates, there are other structures such as the National Agency for Environmental, Food, Occupational and Health Product Safety (ANSSEAT), whose mission is just as important in the fight against and prevention of NCDs in Burkina Faso, but no functional link between the DPCM and ANSSEAT was noted by the actors interviewed.

**Figure 2.**
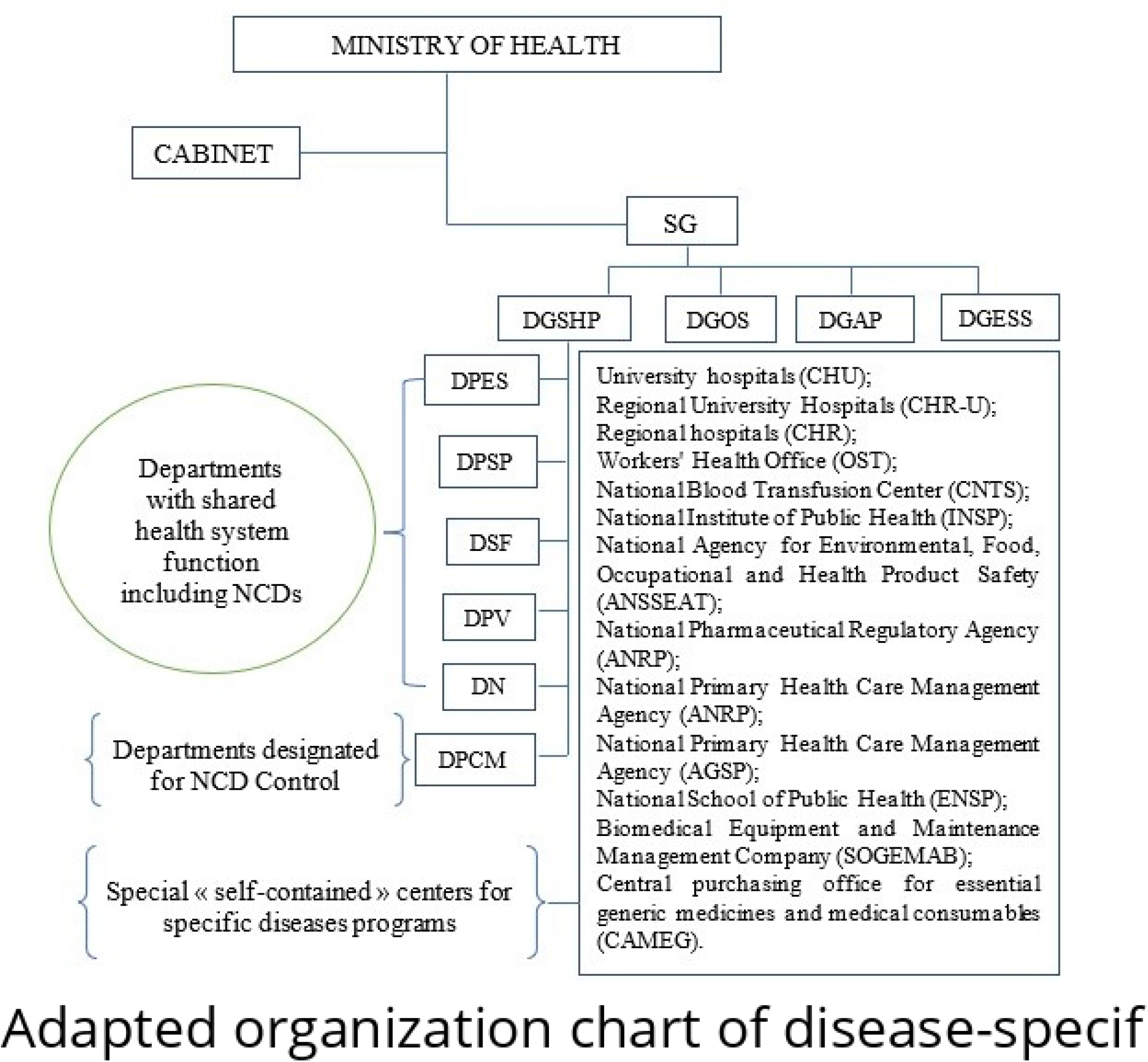
Adapted organization chart of disease-specific structures and overall health function structures within Burkina Faso’s Ministry of Health.

### 3.2. Reference documents

The document review and interviews conducted indicate the existence of several reference documents related to the fight and control of food-related NCDs in several different service directorates. In 2016, the DPCM, which is also the structure in charge of the prevention and control of NCDs, drew up an integrated strategic plan to combat NCDs (2016 - 2020), budgeted at 5,469,734,500 FCFA or around 9.8 million dollars ($). In addition to this integrated strategic plan, the DPCM has other plans in place, notably (i) the three-year oral health strategic plan, (ii) the strategic action plan to combat cancer, all of which are currently being implemented in addition to the integrated strategic plan, which expires in 2020. It should be noted that the integrated strategic plan to combat NCDs has not been evaluated either at mid-term or at the end of its implementation, although the DPCM has initiated a review of this reference document. Although all the players interviewed recognize the need to revise the plan to adapt it to the new requirements in terms of control and fight against NTMs, they are unanimous (100% of those interviewed) that an evaluation of the current plan should have been carried out to identify shortcomings and bottlenecks, in order to correct them in the new reference system currently being drawn up. “*The development of such a document should be based on evidence and new WHO guidelines unfortunately the implementation of the plan has not been evaluated to assess the strengths and weaknesses of the current plan prior to the update”*. In addition to these documents, Burkina Faso has taken a number of measures to combat and control NCDs, including (i) a smoking ban, (ii) a ban on tobacco advertising, promotion and sponsorship, (iii) restrictions on the physical availability of alcohol, (iv) an increase in alcohol excise duties, (v) restrictions on the marketing of breast-milk substitutes, (vi) a strong measure on cancer treatment.

### 3.3. Involving stakeholders in the design process

Sixty percent (60%) of those interviewed said they had been involved in drawing up the integrated plan to combat NCDs. It should be noted that certain technical structures of the Ministry of Health declared that they had not been involved in drawing up the plan.

Forty percent (40%) appear not to have been involved, especially NGOs/Associations. “*the drafting of the document was not as participatory; it was mainly Ministry of Health players and NGOs/Associations working in the field of NCDs that were involved”, stresses one of the interviewees”*.

> “…*other ministerial departments such as agriculture, education, trade, water, hygiene and sanitation have not been involved in the process, even though they have a role to play, so I don’t think they feel concerned, even though everyone has a role to play”*.

### 3.4. Existence of a coordinating body or mechanism

All those interviewed (100%) felt that the Ministry of Health was leading the fight against NCDs in Burkina Faso. Analysis of the actors involved in implementing the integrated strategic plan to combat NCDs suggests that planning has been multi-sectoral. Of the 32 actors identified, other ministerial departments other than the Ministry of Health accounted for 28%, the academic sector (9%), the private sector (6%) and United Nations agencies.

As far as multi-sectoral coordination or coordination mechanisms are concerned, no bodies have been set up at either sectoral (Ministry of Health) or national level. All of those interviewed (100%) also deplored the absence of a framework for consultation between these actors to ensure better coordination of the implementation of these interventions. As one informant put it: “*the fight against NCDs is multi-sectoral and cannot be dealt with by the health department alone. The problem is that the roles and responsibilities of each player involved are inadequately defined*… *the other aspect is that we don’t have a framework for concerted action, as in the case of the fight against malnutrition*”.

### 3.5. Resource availability

The interviewees (100%) clearly mentioned the inadequacy of resources allocated to the fight against NCDs in Burkina Faso, both in the state budget and from technical and financial partners:

> *“…most of the interventions set out in the integrated strategic plan to combat NCDs have not been implemented, due to insufficient or even non-existent resources to implement them”*,
>
> *“… it’s as if all the efforts of both the state and its partners are directed towards combating malnutrition, yet the burden of these NCDs is just as real for our populations”*.

For them, there isn’t enough enthusiasm at international level for raising funds to fight NCDs. In support of this view, one interviewee emphasized: *“… for example, every year, Burkina Faso carries out nutritional surveys with the support of technical and financial partners to assess the nutritional status of children under 5 years of age. However, in 10 years, Burkina Faso has only carried out two (2) STEPS surveys, which shows how much interest people have in the problems of NCDs in our country»*. The interviewee sees this as a clear indicator of the lack of commitment and interest in the NCD issue in developing countries in general, and in Burkina Faso in particular.

### 3.6. Identified budget lines

A total of twenty-nine (29) budget lines related to the prevention and/or management of NCDs in Burkina Faso were identified. Following categorization and weighting, three (3) budget lines were considered “specific” to NCDs, eighteen (18) were “sensitive” and eight (8) were considered “positive investments” (Table 1).

**Table I:**
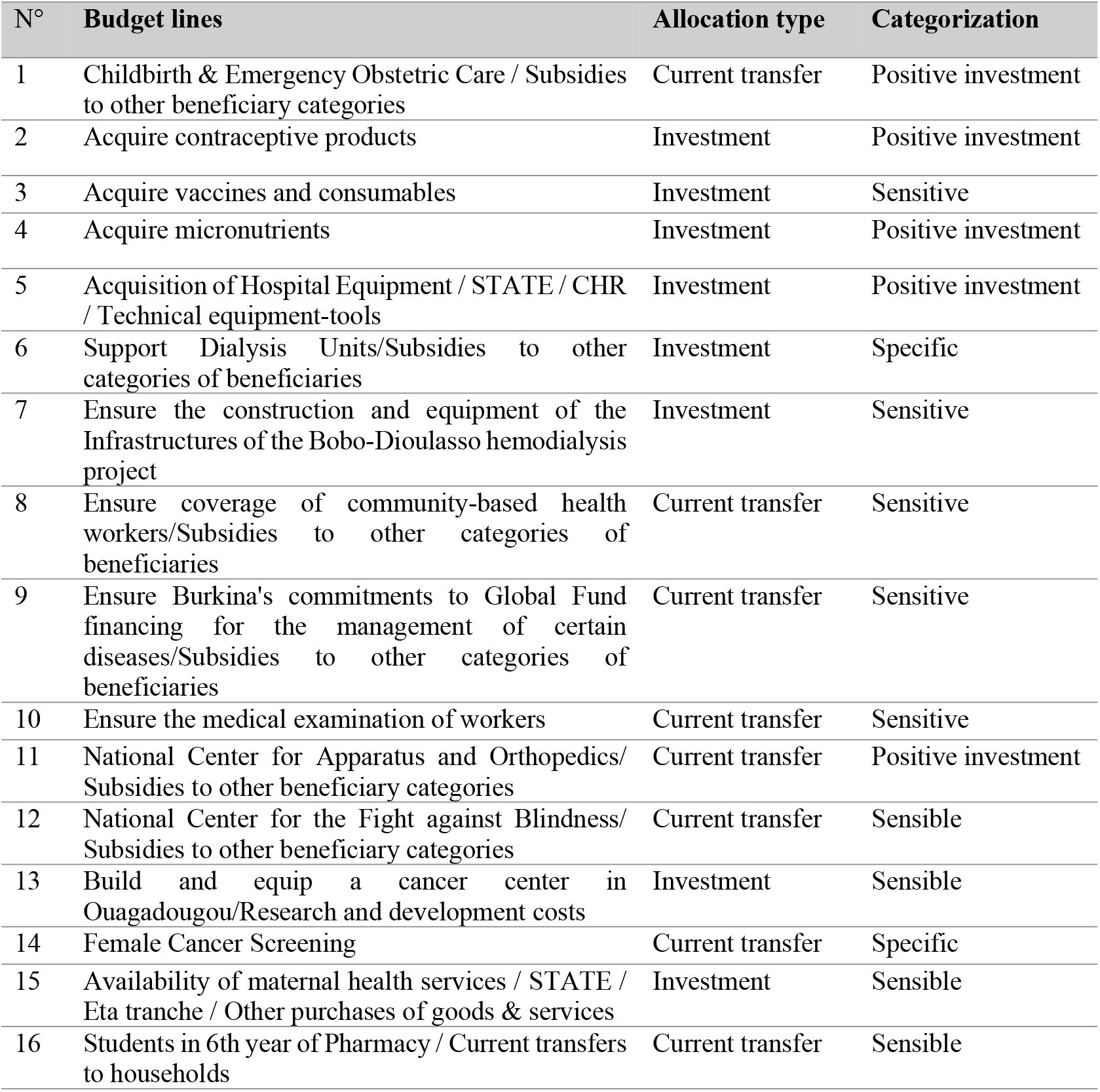
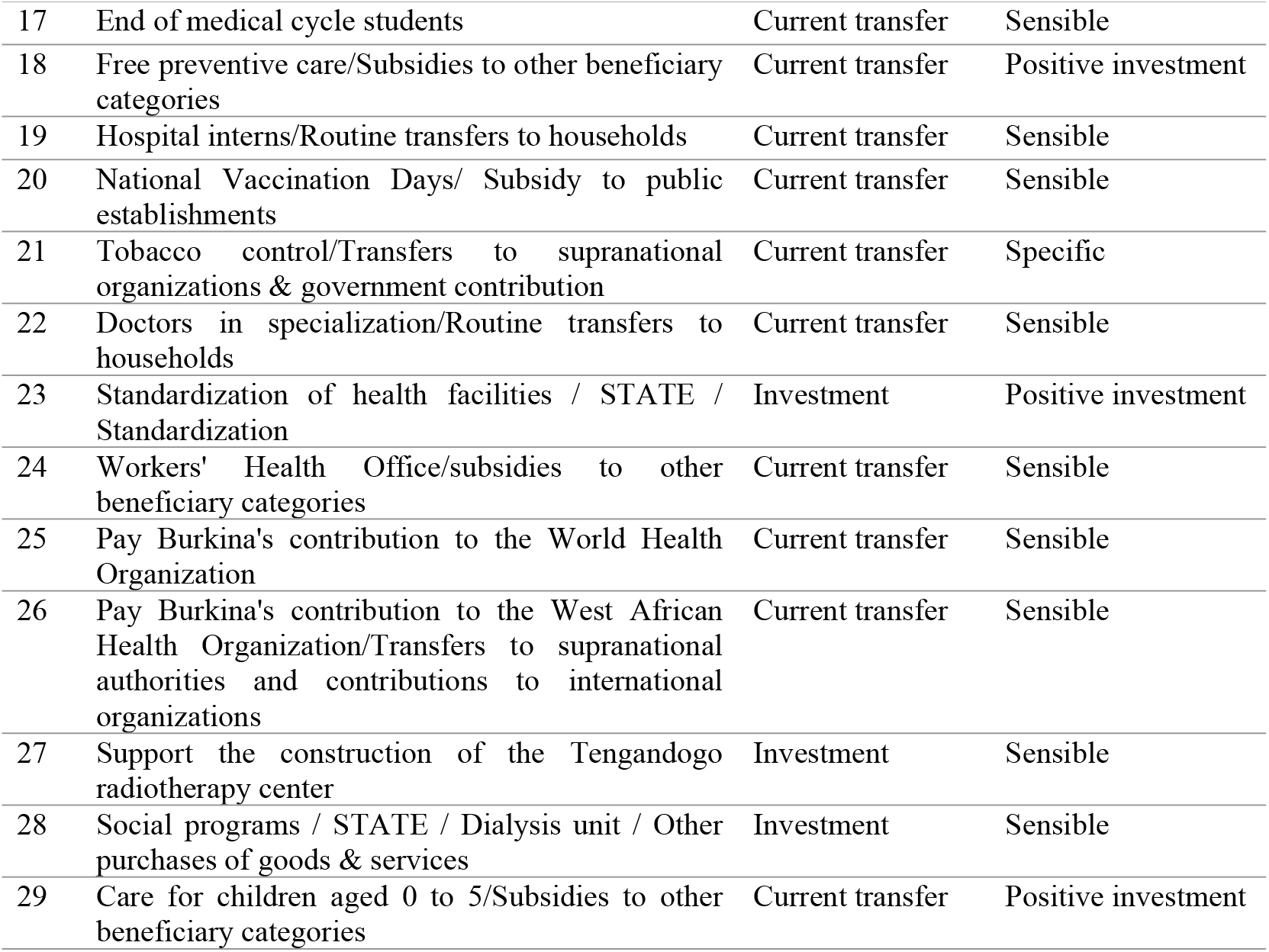
Selected budget lines.

| N° | Budget lines | Allocation type | Categorization |
| --- | --- | --- | --- |
| 1 | Childbirth & Emergency Obstetric Care / Subsidies to other beneficiary categories | Current transfer | Positive investment |
| 2 | Acquire contraceptive products | Investment | Positive investment |
| 3 | Acquire vaccines and consumables | Investment | Sensitive |
| 4 | Acquire micronutrients | Investment | Positive investment |
| 5 | Acquisition of Hospital Equipment / STATE / CHR / Technical equipment-tools | Investment | Positive investment |
| 6 | Support Dialysis Units/Subsidies to other categories of beneficiaries | Investment | Specific |
| 7 | Ensure the construction and equipment of the Infrastructures of the Bobo-Dioulasso hemodialysis project | Investment | Sensitive |
| 8 | Ensure coverage of community-based health workers/Subsidies to other categories of beneficiaries | Current transfer | Sensitive |
| 9 | Ensure Burkina's commitments to Global Fund financing for the management of certain diseases/Subsidies to other categories of beneficiaries | Current transfer | Sensitive |
| 10 | Ensure the medical examination of workers | Current transfer | Sensitive |
| 11 | National Center for Apparatus and Orthopedics/ Subsidies to other beneficiary categories | Current transfer | Positive investment |
| 12 | National Center for the Fight against Blindness/ Subsidies to other beneficiary categories | Current transfer | Sensible |
| 13 | Build and equip a cancer center in Ouagadougou/Research and development costs | Investment | Sensible |
| 14 | Female Cancer Screening | Current transfer | Specific |
| 15 | Availability of maternal health services / STATE / Eta tranche / Other purchases of goods & services | Investment | Sensible |
| 16 | Students in 6th year of Pharmacy / Current transfers to households | Current transfer | Sensible |
| 17 | End of medical cycle students | Current transfer | Sensible |
| 18 | Free preventive care/Subsidies to other beneficiary categories | Current transfer | Positive investment |
| 19 | Hospital interns/Routine transfers to households | Current transfer | Sensible |
| 20 | National Vaccination Days/ Subsidy to public establishments | Current transfer | Sensible |
| 21 | Tobacco control/Transfers to supranational organizations & government contribution | Current transfer | Specific |
| 22 | Doctors in specialization/Routine transfers to households | Current transfer | Sensible |
| 23 | Standardization of health facilities / STATE / Standardization | Investment | Positive investment |
| 24 | Workers' Health Office/subsidies to other beneficiary categories | Current transfer | Sensible |
| 25 | Pay Burkina's contribution to the World Health Organization | Current transfer | Sensible |
| 26 | Pay Burkina's contribution to the West African Health Organization/Transfers to supranational authorities and contributions to international organizations | Current transfer | Sensible |
| 27 | Support the construction of the Tengandogo radiotherapy center | Investment | Sensible |
| 28 | Social programs / STATE / Dialysis unit / Other purchases of goods & services | Investment | Sensible |
| 29 | Care for children aged 0 to 5/Subsidies to other beneficiary categories | Current transfer | Positive investment |

### 3.7. Budget allocations and expenditure

Over the period 2010 to 2020, the Burkina Faso Ministry of Health allocated nearly 17.33 billion FCFA (US$29.9 million), or an average of US$2.72 million per year, to the fight against NCDs. This allocation represents around 1.55% of the total budget of the Ministry of Health over the same period. The budget absorption rate, defined as the percentage of the allocated budget actually used, was over 98% (Table 2). With this high absorption rate, we carried out the analyses with budget allocations only.

**Table II:** Budget allocations and expenditure related to NCDs of the Ministry in charge of Health from 2010-2020.

| Year (2010 – 2020) | Total | Yearly average |
| --- | --- | --- |
| Total Budget allocations for NCDs (FCFA) | 29 885 033,32<sup>1</sup> \$ | 2 716 821 \$ |
| Total Expenses for NCDs (FCFA) | 29 459 619,47 \$ | 2 678 147,22 \$ |
| Absorption rate (expenses / allocations in %) | 98,58% | 98,58% |

<sup>1</sup> 1 dollar US = 580 FCFA

### 3.8. Allocation by intervention type

Of this budget of 17.33 billion FCFA ($29.8 million), 6.64 billion or 38.4% was allocated to “specific NTM” interventions, while 10.68 billion or 61.6% was allocated to “sensitive” interventions.

### 3.9. Budget allocation by objectives of the global action plan

Analysis of the budget according to the objectives of the WHO global action plan to combat NCDs 2013-2020 suggests that only interventions aimed at achieving 3 of the 6 WHO objectives, namely objectives 3, 4 and 5, received a budget allocation. Goal 4, aimed at strengthening and orienting health systems, received the largest share of the allocation, i.e. 13.6 billion or 78.48% of total funding. Objective 5, which aims to promote and strengthen national capacity to carry out actions to prevent and combat NCDs, was the second most financed with 2.08 billion, or around 12%, and finally objective 3, which aims to reduce exposure to modifiable risk factors, received 1.64 billion, or 9.5%.

The objectives of the action plan that received no budgetary allocation include Objective 1 “Give greater priority to the fight against NCDs in global, regional and national agendas and in internationally agreed development goals, by strengthening international cooperation and awareness”, Goal 2 “Strengthen national capacity, leadership, governance, multisectoral action and partnerships to accelerate action on NCDs in countries” and Goal 6 “Monitor trends and determinants of NCDs and assess progress in prevention and control”.

### 3.10. Evolution of budget allocations from 2010 – 2020

Trend analysis of allocations shows an upward trend in annual allocations from 365 million in 2010 to over 5 billion in 2017, with a rollercoaster ride between 2010 and 2013 (Figure 3). Allocations then underwent a drastic reduction of over 62%, from 5.019 billion in 2017 to 1.877 billion in 2020, virtually their 2016 value.

**Figure 3.**
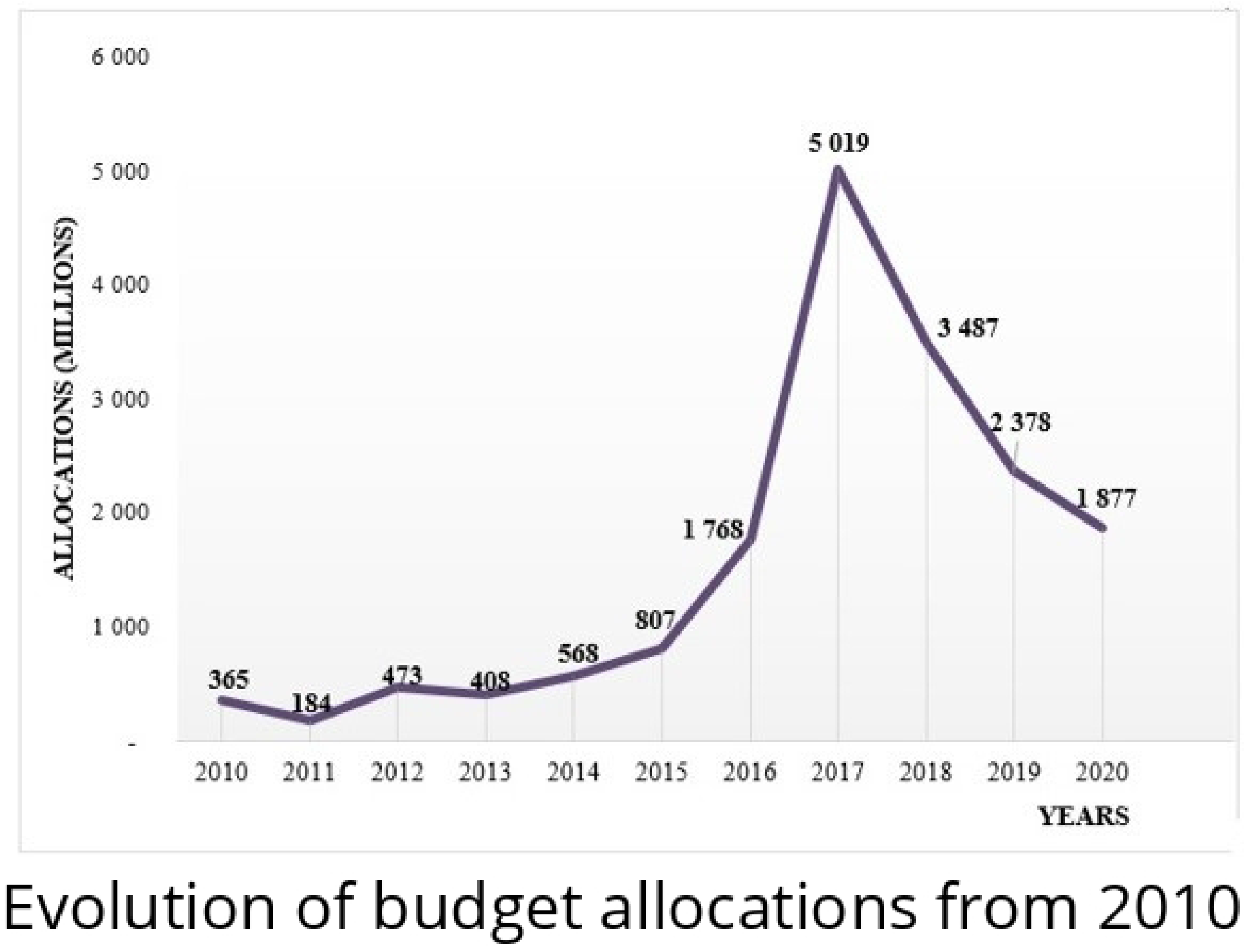
Evolution of budget allocations from 2010 – 2020

### 3.11. Allocations before and after the development of the integrated strategic plan for the fight against NCDs 2016-2020

In 2015, the Ministry of Health drew up an integrated national strategic plan to combat NCDs 2016-2020. Immediately after the adoption of this integrated strategic plan, the annual budget dedicated to NCDs more than doubled, from 807 million CFA francs in 2015 to 1.87 billion CFA francs in 2020 (Figure 4). Funding peaked at 5.019 billion in 2017, representing a 522% increase on 2015, the year before the National Strategic Plan 2016-2020 came into force.

**Figure 4.**
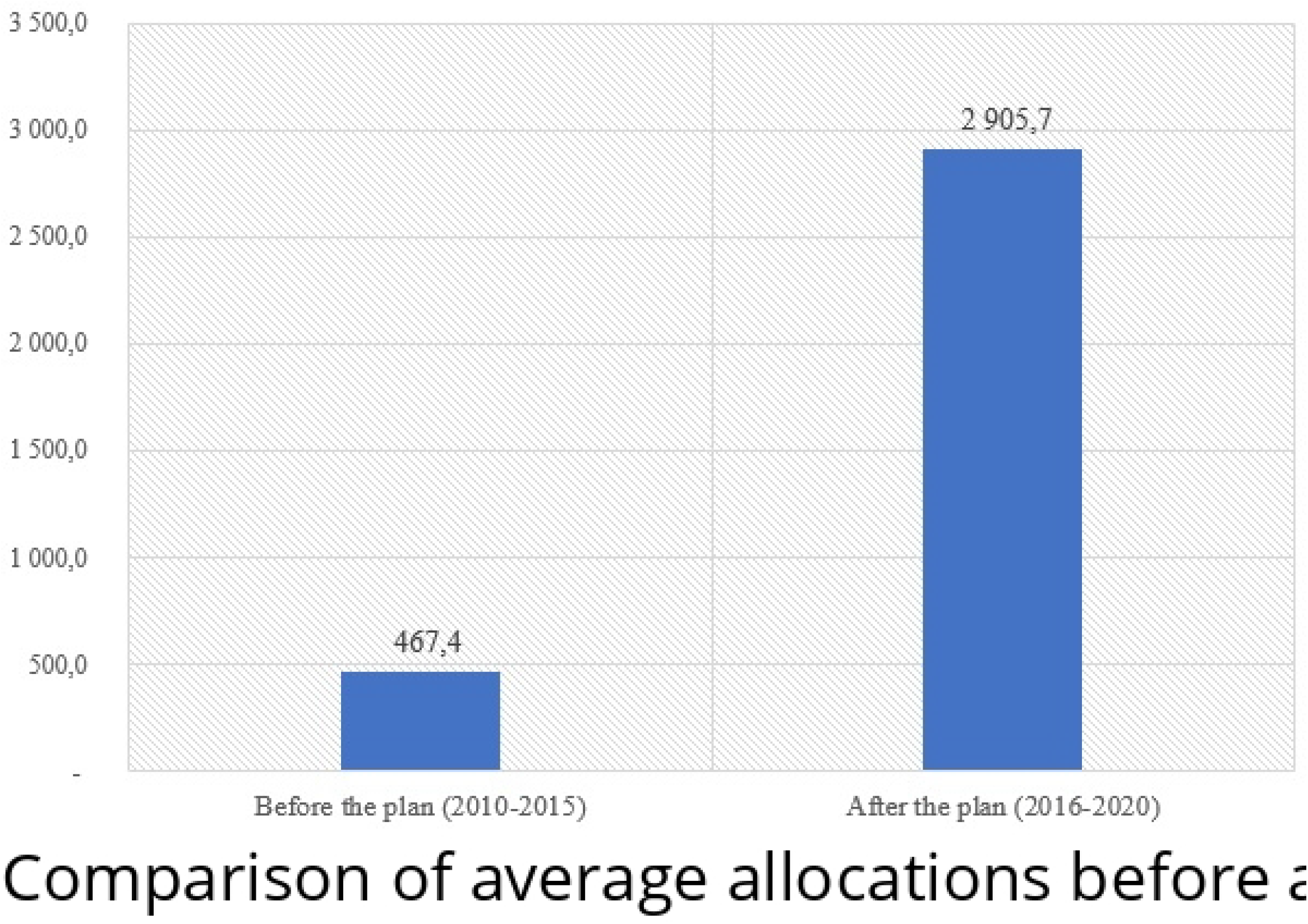
Comparison of average allocations before and after the strategic plan

A comparison of average annual budget allocations before (2010-2015) and after (2016-2020) the development of the strategic plan (Figure 4) shows that, overall, budget allocations improved after the strategic plan was developed. Indeed, the average annual allocation increased significantly (p=0.02) from around 467.4 million before the adoption of the strategic plan to over 2.9 billion FCFA afterwards.

## 4. Discussions

This study provides an answer to one of the concerns raised by Ouedraogo et al, [14] about institutional arrangements and budget allocations for the prevention and control of NCDs in Burkina Faso.

Burkina Faso has drawn up a number of reference documents relating to the fight against NCDs, notably the integrated plan to combat NCDs (2016-2020). This document was drawn up at the level of the Ministry of Health and did not take sufficient account of other ministerial departments, notably agriculture, education, water, hygiene and sanitation, and the private sector, despite the fact that the multi-sectoral nature of the fight against food-related NCDs has been documented by several authors [24, 25]. In Morocco and Algeria, for example, the title of the reference document is very evocative. Indeed, Morocco has a national multisectoral strategy for the prevention and control of non-communicable diseases and Algeria has a national multisectoral strategic plan for the integrated fight against the risk factors of non-communicable diseases [26, 27]. Ghana has a national policy for the prevention and control of NCDs (Ghana NCD Policy 2014) [28]. Such reference documents developed in a multi-sectoral dynamic with a national character allow for commitment at the highest level and better advocacy for the mobilization of resources [29, 30] and helps create an environment conducive to the adoption of healthy behaviors [31]. It is therefore essential that Burkina Faso equips itself as soon as possible with a multisectoral policy or plan to combat and control NCDs; moreover, the WHO estimates that Burkina Faso does not have any documents on this subject [9].

No consultation framework or common results framework that could enable multisectoral action and coordination is available in Burkina Faso for the fight against NCDs, yet this step is a necessity for optimal programming and implementation of interventions with a guarantee of accountability of the actors involved [24, 25, 32]. This situation is not typical of Burkina Faso in the ECOWAS region; Ghana, for example, which has had initiatives to combat NCDs since the 1990s with a national policy to combat NCDs, has not succeeded in establishing an effective multi-sectoral consultation framework [28]. The implementation of such a framework allows in some cases better monitoring and better synergy of action of the partners around the priorities. Some countries seem to have been able to set up this framework of consultation like Morocco for example. This is probably due to the fact that the development of the document was participatory and multisectoral from the base. This provision facilitates multisectoral coordination and allows better mobilization of financial and human resources for the implementation of the policy [6, 30, 33].

The absence of multi-sectoral programming and a multi-sectoral consultation body as observed in Burkina Faso leads to an overlap of numerous functions making the mobilization of resources ineffective [34] in a context where international development aid to meet this burden remains negligible [35]. This observation was clearly made by key informants who unanimously stated that there is an insufficiency of resources allocated to the issue of NCDs in Burkina Faso. They even believe that NCDs do not seem to be a priority for either the state or partners in terms of the resources allocated to the issue. The budget analysis carried out showed that during the period 2010-2020, the Ministry of Health of Burkina Faso allocated only 1.5% of its sectoral budget to the fight against NCDs in Burkina Faso, i.e. approximately 17.33 billion FCFA (29.9 million US dollars), thus justifying the statements of the interviewees.

WHO estimates that annual investment in NCD interventions ranges from less than US$1 per person in low-income countries to US$3 per person in upper-middle-income countries [36]. The most effective NCD interventions for individuals cost US$11.4 billion per year for all low- and middle-income countries [36]. Although we are unable to clearly establish the impact of Burkina Faso’s integrated plan on the increase in allocations observed after the adoption of the plan in 2015, the process of developing the plan has enabled us to better understand the problem, to properly structure and quantify the response, and therefore to clarify investment opportunities [4, 24, 37]. The plan highlighted, for example, the importance of “female cancer screening,” which is considered one of the most cost-effective interventions in the fight against NCDs, with a cost-effectiveness ratio of ≤ $1 according to WHO [22]. The inadequate allocation of resources to combat and control NCDs has been documented by the Ghana Noncommunicable Diseases Alliance which identified funding as a major challenge but acknowledged that data on budget allocations for NCDs in Ghana are unavailable. It is clear that there is a lack of resources allocated to combat NCDs yet low and middle income countries like Burkina Faso can gain $350 billion by 2030 by increasing investments in NCD prevention and treatment [38].

According to WHO, for every dollar invested in scaling up NCD response in these countries, society will see a return of at least $7 in increased employment, productivity and longer lives [38]. Burkina Faso would therefore benefit from investing significantly in the fight against NCDs in view of the ever-increasing burden of these diseases.

To our knowledge, this is the only study in Burkina Faso that has provided some answers to questions about the institutional environment and budgetary allocations intended for the fight against NCDs despite the methodological limitations, namely (i) the insufficiency in taking into account the reference documents of other ministerial departments; (ii) institutional changes in Burkina Faso have led to several changes at the head of state structures, which could impact the responses of interviewees depending on their seniority at the head of the structure; (iii) the consideration only of state allocations in the analysis due to the difficulty of obtaining accurate data on funding from technical and financial partners, (iv) the absence of a common multisectoral results framework listing consensual interventions for the fight against NCDs in Burkina Faso, which makes financial monitoring difficult in other contributing sectors.

## 5. Conclusion

The analysis of the institutional framework and budget allocations for the fight and control of NCDs reveals many difficulties in the institutional system in Burkina Faso. The prevention and control of NCDs must be considered as a system in which appropriate and sustainable investment is required. The analysis shows many difficulties hampering the performance of the fight and control of NCDs in Burkina Faso, including (i) the absence of an accurate estimate of the burden of NCDs, (ii) the absence of a multisectoral policy or strategic plan involving all stakeholders, (iii) the absence of a multisectoral coordination body, (iii) the insufficiency of sufficient financial resources, (iv) the insufficiency of research on the problem of NCDs. Although the institutional framework for combating NCDs in Burkina Faso has shortcomings, it must be recognized that Burkina Faso has made some progress in the fight against NCDs with numerous texts and laws aimed at reducing exposure to modifiable risk factors for non-communicable diseases and the underlying social determinants. In addition, many specific and sensitive budget lines for the fight and control of NCDs have been identified in the Burkinabe state budget despite the difficult security and humanitarian situation that the country has been going through since 2018, thus indicating political will and a favorable environment in a context marked by the scarcity of resources. Much work remains to be done when we know that the scale of the burden of NCDs is high and increasing in low- and middle-income countries such as Burkina Faso.

## Data Availability

No restriction

## 6. Author Contributions

Conceptualization: Moussa Ouedraogo, Dia Sanou, Aly Savadogo

Investigation: Moussa Ouedraogo, Dia Sanou, Mahamadé Goubgou

Formal analysis: Moussa Ouedraogo, Mahamadé Goubgou

Methodology: Moussa Ouedraogo, Mahamadé Goubgou, Tirogo Souleymane, Estelle Aissa Bambara, Aly Savadogo.

Validation: Moussa Ouedraogo, Mahamadé Goubgou, Tirogo Souleymane, Estelle Aissa Bambara, Aly Savadogo.

Writing– original draft: Moussa Ouedraogo.

Writing– review & editing: Moussa Ouedraogo, Mahamadé Goubgou, Tirogo Souleymane, Estelle Aissa Bambara, Aly Savadogo.

## 7. Acknowledgments

The authors express sincere gratitude to all health agencies, study participants and research assistants who supported us through the data collection period.

